# Combined analysis of chromatin accessibility, promoter interactions and whole genome sequencing solved the missing heritability in gastric cancer

**DOI:** 10.1101/2024.11.27.24317972

**Authors:** C. São José, M. Ferreira, L. Cordova, A. Pedro, J. Senz, J. Garcia-Pelaez, S. Lobo, I. Gullo, A. André, P. Kaurah, F. Puntieri, J. Glaser, D. Huntsman, S. Mundlos, K. Schrader, C. Oliveira

**Author notes:** **Correspondence:** Carla Oliveira, BSc, PhD.

## Abstract

Missing heritability in hereditary diffuse gastric cancer (HDGC) ranges from 60 to 90%. These HDGC-like families, despite complying with HDGC clinical criteria, lack *CDH1* and *CTNNA1* actionable germline variants, and are not offered HDGC-targeted life-saving disease prevention measures. Herein, we explored novel HDGC predisposition mechanisms affecting the *CDH1*-regulatory network. We called single-nucleotide (SNV) and copy-number variants (CNV) from 19 HDGC-like probands from whole-genome sequencing data and performed gene-ontology analysis. Chromatin enhancer marks and *CDH1* promoter interactions were evaluated in normal stomach by ChIP-seq, ATAC-seq and 4C-seq, variant causality was assessed by RT-PCR, immunohistochemistry and microsatellite instability (MSI) analysis in tumours. Functional analysis was performed using CRISPR-Cas9, RT-PCR and flow cytometry in cell lines, and enhancer assays using mouse embryos. Within the *CDH1* topologically associating domain (TAD), we found two deletions in Family F4 and F9. F4 carried a heterozygous *CDH3* 20kb-CNV triggering *CDH1* mRNA/protein loss in homozygosity by CRISPR-Cas9 editing, similarly to a *CDH1* coding deletion. This 20kb sequence encloses two hypomorphic tissue-specific regulatory elements (REs), each contributing 50% to *CDH1* expression regulation. F9 carried a heterozygous 39bp-intergenic CNV downstream of *CDH1*, triggering *CDH1* mRNA/protein loss by CRISPR-Cas9. F15, presenting gastric but not colorectal cancer, carried an *MLH1* heterozygous 2.7Kb germline CNV overlapping a stomach-specific RE found by ChIP-seq. The gastric tumour of mixed histology displayed Microsatellite instability (MSI), reduced *MLH1* mRNA and protein, and reduced *CDH1* and E-cadherin protein. CRISPR-Cas9 clones mimicking the *MLH1* heterozygous CNV, triggered loss of *MLH1* and *CDH1*/E-cadherin mRNA and protein, similar to a coding deletion. Beyond the *CDH1* TAD and tumour risk syndrome genes, multiple deletions of stomach accessible chromatin sequences were found in particularly young-affected individuals from additional 6 families. This oligogenic pattern impaired specifically mucin genes and multiple immune-related pathways. Herein, we pinpointed novel mechanisms behind HDGC predisposition. One involves deletions of *CDH1*-REs in the TAD or stomach-specific *CDH1*-REs in the *MLH1* locus. The second involves multiple deletions of stomach REs affecting mucin and immune-related genes, favouring a gastric immune-deficient phenotype. Altogether, by combining stomach-specific chromatin accessibility and promoter interactions with whole genome sequencing, we solved the missing heritability in 47% of HDGC-like families within our cohort.

## Introduction

Missing heritability in hereditary diffuse gastric cancer (HDGC) ranges from 60 to 90%. This corresponds to the proportion of families, that despite complying with clinical criteria for HDGC (Blair et al. 2020), cannot be explained by genetic variants in *bonafide* HDGC-associated genes, *CDH1* (ENSG00000039068) and *CTNNA1* (ENSG00000044115), and are called HDGC-like (Oliveira et al. 2009b; Blair et al. 2020). While 10-40% of HDGC families bear heterozygous coding truncating variants (actionable) in *CDH1* (Oliveira et al. 2009b, 2015), only 2% carry *CTNNA1* truncating variants (Lobo et al. 2021). HDGC-like families remain genetically undiagnosed, even after deep analysis using candidate gene, whole exome and whole genome sequencing (WGS) (Hansford et al. 2015; Oliveira et al. 2015; Tedaldi et al. 2019).

Carriers of actionable variants in *CDH1* have increased risk for early-onset diffuse gastric cancer (DGC) and lobular breast cancer (LBC), the HDGC-associated phenotypes (Guilford et al. 1998; Oliveira et al. 2009a; Garcia-Pelaez et al. 2023). Truncating SNVs in *CTNNA1* have also been associated with DGC predisposition in a small fraction of HDGC families (Majewski et al. 2013; Lobo et al. 2021). Complete *CDH1* and *CTNNA1* loss of function, due to somatic inactivation of the remaining wild-type allele, results in loss of cell adhesion, disruption of epithelial architecture and decreased differentiation, features of signet-ring cells characteristic of HDGC-related tumours (Oliveira et al. 2009c; Benusiglio et al. 2019; Gullo et al. 2020).

Identification of *CDH1* and *CTNNA1* actionable variants in families fulfilling HDGC clinical criteria, triggers cascade genetic testing and intensive surveillance through endoscopy with multiple biopsies, and/or prophylactic gastrectomy in asymptomatic carriers. For asymptomatic *CDH1* female carriers breast intensive surveillance using magnetic resonance imaging (MRI) and computed tomography scan (CT-scan) is recommended, alongside the option of (Blair et al. 2020).

Missing heritability hampers the use of personalized life-saving measures, and may lead to prophylactic surgery in individuals lacking genetic diagnosis, solely based on family history (São José et al. 2023a). HDGC-like families, clinically undistinguishable from *CDH1*-HDGC families, often exhibit germline mono-allelic *CDH1* RNA expression in normal tissues and loss of E-cadherin expression in tumours, despite the lack of germline coding SNVs or CNVs in *CDH1* (Pinheiro et al. 2010). These observations pinpoint a likely causality for *CDH1* and argues towards the presence of undiagnosed germline alterations within noncoding *cis*-regulatory elements (REs), controlling *CDH1* expression and function.

Enhancer-promoter communication within the nucleus tightly regulates cell-type-specific gene expression (Spielmann et al. 2018). REs are instrumental for proper gene expression in time and space and enhancers in particular, play an important role in controlling expression of disease-causing genes (Cova et al. 2023). Their mode of action is usually dose-dependent, redundant, orientation-independent, acting through binding of tissue-specific transcription factors and controlling tissue-specific gene expression (Will et al. 2017; Bordeira-Carriço et al. 2022). Enhancer-promoter communication may occur within large genomic distances, which are brought in close spatial proximity upon 3D-chromatin folding (Bonev and Cavalli 2016; Furlong and Levine 2018). Topologically associating domains (TADs) are the functional units of 3D-chromatin architecture, restricting enhancers’ interaction with their target promoters (Rao et al. 2014; Dixon et al. 2016). CNVs affecting TADs’ structure by affecting the boundary between two separated TADs may trigger gene misexpression patterns and cause disease (Franke et al. 2016; Lupiáñez et al. 2016). Assigning function and pathogenicity to noncoding variants found in enhancers and other REs, in the next-generation sequencing era, remains challenging. The integration of multiple omics technologies and functional models is essential to elucidate the impact of RE perturbation and to establish a link to disease predisposition. Herein, we aim to identify and characterize novel HDGC-causing mechanisms involving REs regulating *CDH1* expression to solve the missing heritability in HDGC-like families, resourcing to WGS, chromatin analysis technologies and functional assays in cells and mice.

## Materials and Methods

### Patient selection

Twenty-one patients from 19 HDGC-like families from various genetic backgrounds were admitted to BC Cancer in British Columbia, Canada and fulfilled the 1999 HDGC clinical criteria (Caldas et al. 1999). Families were re-classified according to (Blair et al. 2020) and HDGC-like families were considered if fulfilled criteria 1 and 2. Gastric cancers with diffuse component, with or without an intestinal component (mixed-type MGC) were considered for criteria 1. All patients tested negative for *CDH1* SNVs and CNVs. The present project was approved by the Ethical Committee of Centro Hospitalar Universitário de São João, with internal references 445-20 and 305-19.

### Whole genome sequencing

Genomic DNA extracted from peripheral blood of 21 patients was sequenced using Illumina platform. Sequencing reads were mapped to the human reference genome (GRCh38) using BWA mem (v.0.7.15). Duplicated reads were marked, and their base quality score was recalibrated with GATK (v4.1.9.0, HTSJDK 2.23.0 and Picard 2.23.3).

### Variant calling

SNVs were called with GATK – HaplotypeCaller tool (v4.1.9.0, HTSJDK 2.23.0 and Picard 2.23.3), and Depp Variant (Docker image: 1.3.0) (Poplin et al. 2018). Called SNVs were filtered according to quality control metrics, read depth (≥8) and genotype quality (≥20), and annotated with VEP (v.107). SNVs were prioritized if called by (1) both callers, from which HaplotypeCaller had high quality (≥200), (2A) Depp Variant, or (2B) HaplotypeCaller with extremely high quality (≥500). CNVs calling was performed with LUMPY (v 0.2.13) (Rausch et al. 2012; Layer et al. 2014), DELLY (v.0.9.1) and GRIDSS (v.2.13.2) (Cameron et al. 2017), and annotated with AnnotSV (v. 2.5.2, Tcl (v. 8.6)) (Geoffroy et al. 2018). Structural variants with ≥1Mb length, inter-chromosomal events and variants lacking ‘PASS’ flag were discarded. CNVs were prioritized based on the following criteria: (Tier 1) called by LUMPY and DELLY with ≥90% overlap, and GRIDSS confirmed breakpoints with ≤4bp difference, (Tier 2A) called by DELLY and GRIDSS confirmed breakpoints with ≤4bp difference, (Tier 2B) called by LUMPY and GRIDSS confirmed breakpoints with ≤4bp difference, and (Tier 2C) LUMPY and DELLY with ≥90% overlap. Rare CNVs within the *CDH1* TAD (hg38: chr16: 68550001-69180000) were analysed to identify potential *CDH1* REs (AF<0.01). Rare coding CNVs and truncating SNVs in genes associated with gastrointestinal syndromes (AF<0.01; **Supplementary Table 1**) were assessed to exclude variants in other cancer-predisposing genes and further identify *CDH1* regulatory elements. For the polygenic approach, ultrarare deletions and truncating SNVs (AF<0.00001) observed in fewer than 6/21 patients (<24% of the cohort) were selected for analysis. Deletions in F4, F9 and F15 were validated by Sanger sequencing and SVs in F4 were validated by Pacbio sequencing as well.

### Chromatin availability data

ATAC-seq (ENCSR337UIU, ENCSR970UNF) and ChIP-seq (ENCSR492BHN, ENCSR009RJD, ENCSR751BHO, ENCSR313DUH, ENCSR574USP, ENCSR078LIZ) data from normal stomach and colonic mucosa was collected from phase 3 ENCODE project (Abascal et al. 2020). GeneHancer enhancers and eQTLs were collected from the GeneHancer project (Fishilevich et al. 2017) and cCREs were collected from the ENCODE database (Moore et al. 2020).

### 4C-seq

Fresh gastric tissue from bariatric surgeries was collected and washed in HBSS 1x (Gibco). Gastric mucosa cells were scraped and enzymatically dissociated with collagenase type I (Merck), dispase (Merck), trypsin inhibitor (Sigma), BSA (Nzytech), dithiothreitol (Invitrogen) and HBSS 1x (Gibco) at 37°C and 150rpm for at least 1h. 4C-seq libraries were generated, as previously described (van de Werken et al. 2012; Krijger et al. 2020), containing 1x10^7^ cells, cross-linked in 2% paraformaldehyde and lysed. Nuclei suspensions were digested with DnpII (New England Biolabs) as primary and Csp6I (New England Biolabs) as secondary restriction enzymes and re-circularized with T4 DNA Ligase (Thermo Fisher Scientific). 4C-seq libraries were purified using Amicon Ultra-15 10 kDa (MWCO) (Millipore) and PCR-amplified with 3.2µg per reaction (**supplementary table 2**). Samples were paired-end sequenced on Illumina HiSeqX technology (150bp reads) according to standard protocols. *CDH1* interactions on a genome scale were mapped from sequenced 4C libraries, using a bioinformatics pipeline based on Pipe4C (Krijger et al. 2020), PeakC for *cis* interactions (Geeven et al. 2018) with window size 2, alpha fdr 0.1 and minimal distance 500 and fourSig for *trans* interactions with window size 5, 1000 iterations, 0.001 fdr and 0.01 fdr probability and considered interactions in categories 1 and 2.

### Immunohistochemistry (IHC)

Paraffin-embedded blocks containing adequate representation of the advanced tumour were selected for IHC testing. Parallel sections 5 μm thick were cut and stained with E-cadherin and *MLH1* (1:150; clone G168-728, BD Biosciences), or P-cadherin, using standard automated techniques. For each antibody, positive and negative external controls were placed on the same slide or on a separate control slide. Hematoxylin-eosin– and IHC-stained slides were analysed by two experienced pathologists.

### gDNA and RNA paraffin extraction

Tumour/normal tissue was marked by an experienced pathologist and gDNA/RNA was extracted from six 10 μm thick slides. Tumour areas with at least 75% tumour cells were selected for extraction, using MagMax FFPE DNA/RNA Ultra Kit (Applied Biosystems, Life technologies), according to the manufacturer’s instructions. Briefly, sections of paraffin-embedded tumour samples were de-paraffinized and protease K-digested. DNA and RNA were bonded to magnetic beads, pelleted against a magnetic stand, and the supernatant containing DNA/RNA washed and eluted.

### Microsatellites instability analysis (MSI)

Tumour tissue of the *MLH1* proband was studied for MSI using a panel of at least five dinucleotide repeat sequences, as described previously (Velho et al. 2008). Tumour was classified as 1) MSI-H if presented instability in ≥ 40% of the markers, 2) MSI-L if presented instability in <40% of the markers and 3) MSS if showed no instability in the tested markers.

### DNA Methylation Analysis

gDNA extracted from tumour and normal counterpart were treated with bisulfite using Epitect Bisulfite kit (Qiagen). *MLH1* promoter was amplified by PCR using Multiplex PCR kit (Qiagen) and the primers depicted in **supplementary table 3**. PCR products were analysed in gel electrophoresis and Sanger sequenced on an ABI-3130 Genetic Analyzer (Applied Biosystems).

### Cell lines and culture conditions

Human cell line MKN74 was purchased from the JCRB Cell Bank. MKN74 cell line and isogenic clones were cultured in RPMI medium (Gibco) supplemented with 10% foetal bovine serum (Biowest) and 1% Penicillin-Streptomycin (Gibco). HEK293T was cultured in DMEM medium supplemented with 10% foetal bovine serum (Biowest) and 1% Penicillin-Streptomycin (Gibco). Cells were maintained at 37°C and 5% CO_2_ in a high humidity atmosphere. Cell identification was confirmed by STR analysis and cells were confirmed to be free of mycoplasma contamination.

### In vitro functional assay by CRISPR-Cas9

To test enhancer function, each region was targeted with CRISPR-Cas9. sgRNAs were designed using Benchling online platform (**supplementary table 3**). Individual sgRNAs (Invitrogen) were cloned in LentiCRISPRv2GFP (addgene 82416) or LentiCRISPRv2-mCherry (addgene 99154) vectors using BsmBIv2 (New England Biolabs). Plasmids were transformed into Stbl3 competent cells and colonies were sequenced (**supplementary table 3**). Lentiviral particles were produced resourcing to HEK293T cell line with pMD2.G (addgene 12259) and pCMV-dR8.91 (addgene) vectors, following Lipofectamine 3000 manufacture’s protocol (Invitrogen) and collected at 48h. MKN74 was infected with pairs of lentivirus particles in medium supplemented with 10μg/μl hexadimethrine bromide (Merck Life Science S.L.U.) for 48h. Transduced cells were selected for GFP and mCherry positive expression at 7 days post-infection using FACS ARIA (BD Biosciences).

### Genotyping of edited clones

gDNA was extracted using NZY Tissue gDNA isolation kit (NZYTech), according to the manufacturers’ protocol. gDNA was amplified using primers flanking the edition sites (**supplementary table 3**) and Multiplex PCR kit (Qiagen). PCR products were analysed in gel electrophoresis and Sanger sequenced on an ABI-3130 Genetic Analyzer (Applied Biosystems).

### CDH1/E-cadherin and CDH3/P-cadherin expression analysis

*CDH1*/*3* and *MLH1* mRNA expression was assessed by qPCR in triplicates for the MKN74 isogenic clones and in a single experiment for tumour/normal tissues from probands. RNA was extracted using mirVana RNA Isolation Kit (Invitrogen), according to manufacturers’ protocol. cDNA was synthesized using SuperScriptII reverse transcriptase (Invitrogen), according to the manufacturers’ protocol. mRNA expression was analyzed by qPCR with KAPA PROBE FAST qPCR Master Mix (2X) Kit (Sigma-Aldrich) and probes for *CDH1* (Hs.PT.58.3324071, TaqMan), *CDH3* (Hs.PT.51.5028751, IDT), *MLH1* (custom assay, IDT) and 18S (custom assay, IDT). Reactions were sequenced on a 7500 Real-Time PCR System (Applied Biosystems). Relative expression was normalized for the endogenous 18S control and quantified using the 2^−ΔΔCt^ method.

E-cadherin, P-cadherin and *MLH1* expression was assessed by flow cytometry in triplicates. Cells were detached with Versene (Gibco) and blocked with 3% bovine serum albumin-phosphatase buffer saline (Gibco). Cells were incubated with primary mouse monoclonal antibody HECD-1, P-cadherin or *MLH1* (1:100 dilution; 1h at 4°C; Invitrogen), washed and incubated with secondary antibody anti-mouse Alexa Fluor 647 (Invitrogen). Fluorescence was measured using FACS Fortessa or Canto II (BD Biosciences) and Flow Jo version 10 software was used to analyse the data.

### Statistical analysis

Statistical analysis was performed using GraphPad Prism version 7.00 software (GraphPad Software Inc.). A t-student test was used for comparison analysis, assuming equal variance between clones and parental samples. Differences were considered significant when p-value<0.05.

### Enhancer-reporter assays in vivo

Enhancer activity was analyzed in transgenic mouse embryos using a site-specific integration protocol adapted for mouse embryonic stem cells (mESCs), as described previously (Phan et al. 2024). Briefly, the PhiC31 system (Chi et al. 2019) facilitated precise recombination between two att sites: an attP site inserted in a landing pad at the H11 safe-harbor genomic locus and an attB site within a donor vector containing the candidate enhancer and a puromycin selection marker (Sigma-Aldrich, P8833).

First, a mESC line was generated by CRISPR/Cas9 (FuGENE technology, Promega) to insert the H11 landing pad, which contained the Hsp68 promoter upstream of the LacZ reporter gene (Hsp68::LacZ). This cell line served as a negative control to assess background staining. Candidate enhancer regions were PCR-amplified from human genomic DNA using PrimeSTAR GXL DNA Polymerase (Takara Bio) and primers with appropriate overhangs (**supplementary table 4**). Donor vectors were cloned by Gibson assembly (New England Biolabs; (Gibson et al. 2009)).

Subsequently, each donor plasmid was co-transfected with a PhiC31 plasmid into the Hsp68::LacZ mESC line using Lipofectamine LTX (Invitrogen). Transfected cells were selected with puromycin, and genetically modified mESCs were used to generate transgenic embryos via morula aggregation (Artus and Hadjantonakis 2011). CD1 female mice served as foster mothers.

At embryonic day E14.5, the embryos were harvested and processed for beta-galactosidase activity. Embryonic tissues were dissected in iced-cold 1X PBS, fixed in 4% paraformaldehyde (PFA) for 10 minutes and stained in beta-galactosidase solution at 37°C for a several hours to overnight, following Lobe et al. (1999). Dissected organs were imaged using a ZEISS SteREO Discovery.V12 microscope with a cold light source CL9000 and a Leica DFC420 digital camera (Leica Microsystems). Embryos were stored in 4% PFA in 1x PBS at 4°C.

## Results

We whole genome sequenced blood gDNA from 21 HDGC-like patients. Description of the cohort is depicted in **supplementary figure 1**. Following variant calling, we prioritized rare SNV, CNV and SV (AF<0.01), based on low frequency in cancer-free individuals in the *CDH1* TAD and in cancer syndrome-associated genes (**supplementary table 1**), and ultrarare deletions and truncating SNVs (AF<0.00001) that were common in our cohort (<6 patients, <24%).

### Variants within REs in the CDH1 TAD solve missing heritability in two HDGC families

We identified potential REs within the *CDH1* TAD by extracting biochemical and chromatin annotations, such as histone modifications from ChIP-seq data in normal stomach. These marks were overlapped with regions that physically interact with the *CDH1* promoter in the same tissue, obtained through 4C-seq, and that display accessible chromatin identified through ATAC-seq. The list of potential REs was then crossed with CNVs and SVs found in HDGC families through WGS. We found a heterozygous deletion encompassing *CDH3* (encoding the P-cadherin protein) exons 1 and 2, intron 1 and part of intron 2, as well as two inversions (SVs) overlapping each side of the deletion boundaries in the proband of family F4 (**figure 1A,B**). These structural variants, validated by Pacbio long-read sequencing and Sanger sequencing, revealed a 20kb-deleted allele (a), and two inversions flanking the deletion, retaining most of the deleted sequence (b). However, the 5’ and 3’ breakpoints of this allele were not determined (b) (**figure 1B**). In addition, we found evidence for a wild-type sequence including the deleted region with non-inverted flanking regions (c) (**figure 1B**).

**Figure 1.**
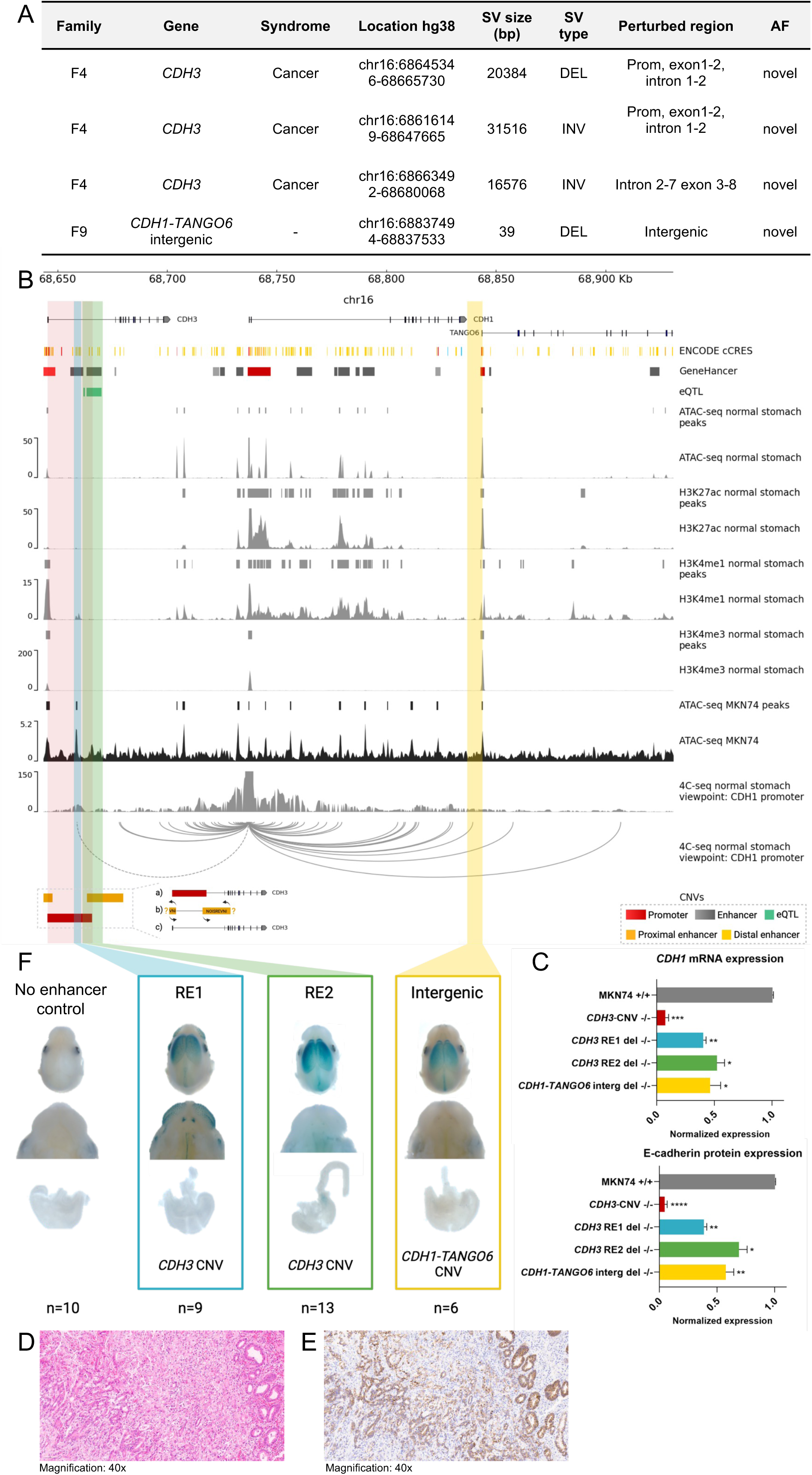
Genomic CNVs in CDH1 TAD. A) Description of identified CNVs, B) Candidate REs, chromatin marks, and CDH1 interactors in normal stomach tissue, C) CDH1 mRNA and protein expression of edited MKN74 clones, D) Hematoxylin and eosin of tumour tissue from proband with CDH3 CNV, E) E-cadherin expression of tumour tissue from proband with CDH3 CNV (immunohistochemistry), F) Enhancer assay of candidate REs in mouse embryos, blue precipitate represent β-galactosidase expression and tissue-specific enhancer activity. GeneHancer: gradient of red and grey represent promoters and enhancers, respectively, grading from low to high activity.

We focused on the *CDH3*-CNV impact in *CDH1* expression, by CRISPR-Cas9 deleting the *CDH3* 20kb sequence in a stomach-derived cell line. The homozygous deletion of this region led to complete loss of *CDH1* mRNA and E-cadherin protein (**figure 1C**), similar to a *CDH1* coding deletion (São José et al. 2023b). The homozygous *CDH3-*CNV also triggered complete loss of *CDH3*/P-cadherin expression (**supplementary figure 3 A, B**), as the deleted sequence includes the *CDH3* promoter, exon 1 and exon 2 (**figure 1A**). The tumour from family F4 proband presented mixed-type histology with signet ring cells, characteristic of HDGC tumours, and E-cadherin expression loss (**figure 1D, E**). Altogether, these results suggest that *CDH1* expression is regulated by REs lying within the *CDH3* 20kb deleted sequence.

As dramatic expression changes often depend on multiple REs being perturbed, we looked for REs within the 20kb deleted region. We found two potential *CDH1* REs within the deletion, namely a sequence interacting with the *CDH1* promoter in normal stomach epithelia found by 4C-seq, that displays accessible chromatin in a gastric cell line with normal *CDH1* expression (RE1), and a second sequence encompassing a *CDH1*-controlling expression quantitative trait loci (eQTL, RE2) (Fishilevich et al. 2017) (**figure 1B**). CRISPR-Cas9 homozygous deletion of RE1 alone led to 60% expression loss in *CDH1* mRNA and E-cadherin protein levels, while RE2 homozygous deletion alone caused 40% expression loss in *CDH1* mRNA and E-cadherin protein levels. The sum of the perturbation of RE1 and RE2 independently, mimicked the impact of deleting the complete 20kb sequence (**figure 1C**). Deletion of the intronic RE1 did not interfere with *CDH3* mRNA nor with P-cadherin protein expression, while deletion of the intronic RE2 led to 30% expression loss in *CDH3*/P-cadherin (**supplementary figure 3 A,B**). These data suggest that RE2 modulated both *CDH1* and *CDH3* expression, while RE1 modulated *CDH1* expression only. Enhancer reporter assays in mouse embryos revealed strong enhancer activity in the forebrain for RE1 and RE2 sequences (**figure 1F**). While RE1 showed strong enhancer activity in the closing palate, teeth, whiskers and eye, RE2 had mild activity in the stomach (**figure 1F**), correlating well with tissues known to express *CDH1* during development (Stemmler et al. 2003). Inversion of the 20Kb deleted region in F4, does not impact *CDH1* expression (*data not shown*). As for the allele bearing the two inversions, the 5’-inversion places *CDH3* exon 2 ahead of exon 1 and part of the promoter, which is predicted to disrupt *CDH3* transcription and translation initiation. A similar effect is expected for the 3’-inversion that inverts the region spanning exon 3-8. The impact of these inversions in *CDH1* expression was not assessed, yet the *CDH3*-CNV correlates well with *CDH1* expression loss and HDGC-related phenotypes observed in family F4.

The tumour spectrum of Family F4 is broad, enclosing three gastric cancers (ages <40s), a central nervous system tumour (age 40s), an ovarian cancer (age 20s), a skin cancer (age 50s) and a breast cancer (**figure 2A**). This young age of onset and broader tumour spectrum may derive from the combined perturbation of *CDH1* and *CDH3*, impacting tissues beyond the classically associated with *CDH1* impairment, and the activity of the deleted REs in different tissues, as illustrated by the strong forebrain enhancer activity found in mice and the presence of a central nervous system tumour (**figure 1F, 2A**).

**Figure 2.**
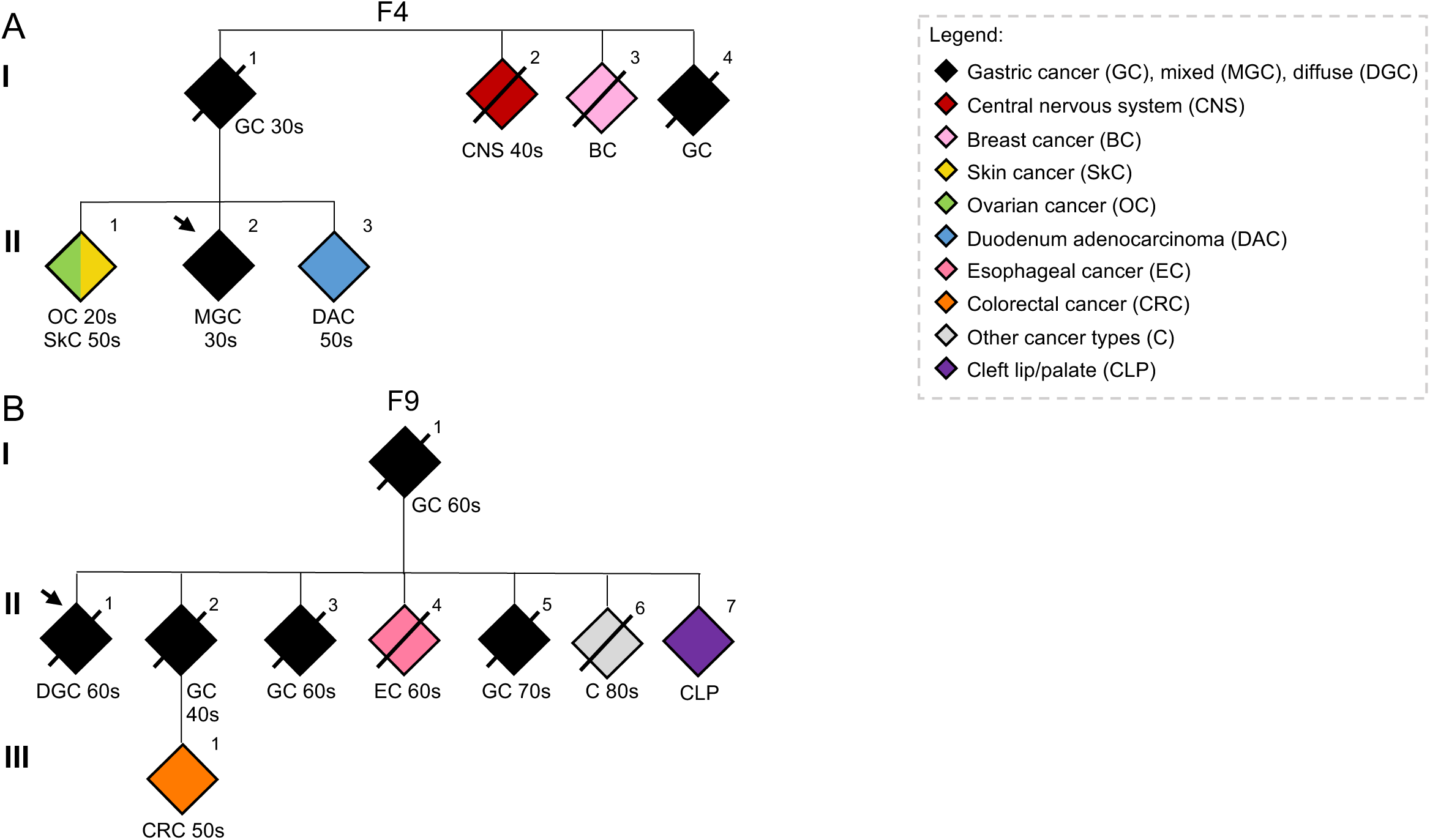
Family pedigrees. A) Representative family pedigree of F4 carrying the CDH3 CNV, B) Representative family pedigree of F9 carrying the CDH1-TANGO6 intergenic CNV. Black fill represents gastric cancer, red represent central nervous system cancer, light blue represents breast cancer, yellow represent skin cancer, green represent ovarian cancer, grey represent endometrial cancer and blue represent colorectal cancer. Arrow indicates the proband. DGC: diffuse gastric cancer, MGC: mixed gastric cancer, GC: gastric cancer, DAC: adenocarcinoma of the duodenum, CNS: central nervous system cancer, BC: breast cancer, SC: skin cancer, OC: ovarian cancer, EC: Esophageal cancer, CRC: colorectal cancer, CLP: cleft lip/palate.

We found that family F9, presenting with four gastric cancers (one in age 40s and three in age 60s), three of which after age 60, and one colorectal cancer in 50s (**figure 2B**), carried a 40bp deletion within the *CDH1-TANGO6* intergenic region, bearing *CDH1*-promotor interactions in normal stomach (**figure 1A, B**). Homozygous deletion of the entire *CDH1-TANGO6* intergenic region led to 50% reduction loss in *CDH1* mRNA and E-cadherin protein levels (**figure 1C**). The deleted region showed enhancer activity in mouse embryos forebrain, but not in the stomach or colon (**figure 1F**). Given that, in homozygosity, this deletion triggers half *CDH1* expression loss, a third event may be required for gastric cancer (GC) development, which can explain the latter age of cancer onset seen in this family.

Altogether, we reveal a novel mechanism impacting *CDH1* expression, through the impairment of hypomorphic and dose-dependent enhancers within the *CDH1* TAD, likely explaining cancer predisposition in two HDGC-like families.

### A variant within a long-distance stomach-specific RE solves missing heritability in one HDGC family

To further solve the missing heritability in the remaining HDGC-like families, we inquired the WGS data from the remaining HDGC-like patients for rare (AF<0.01) SNVs, CNVs and SVs in cancer syndrome-associated genes depicted in **supplementary table 1**. We found a 2.787-bp deletion encompassing the *MLH1* out-of-frame exon 13 and part of introns 12 and 13 in family F15 (**figure 3A**). *MLH1* is a Lynch syndrome mismatch repair gene associated with Lynch syndrome, and its loss in cancer cells leads to microsatellite instability (MSI). Indeed, the F15 proband’s tumour showed *MLH1* expression loss, MSI and lacked *MLH1* promoter methylation (**Figure 3B-D, supplementary figure 3 E**), arguing towards the causality of the *MLH1*-CNV in HDGC predisposition in this family.

**Figure 3.**
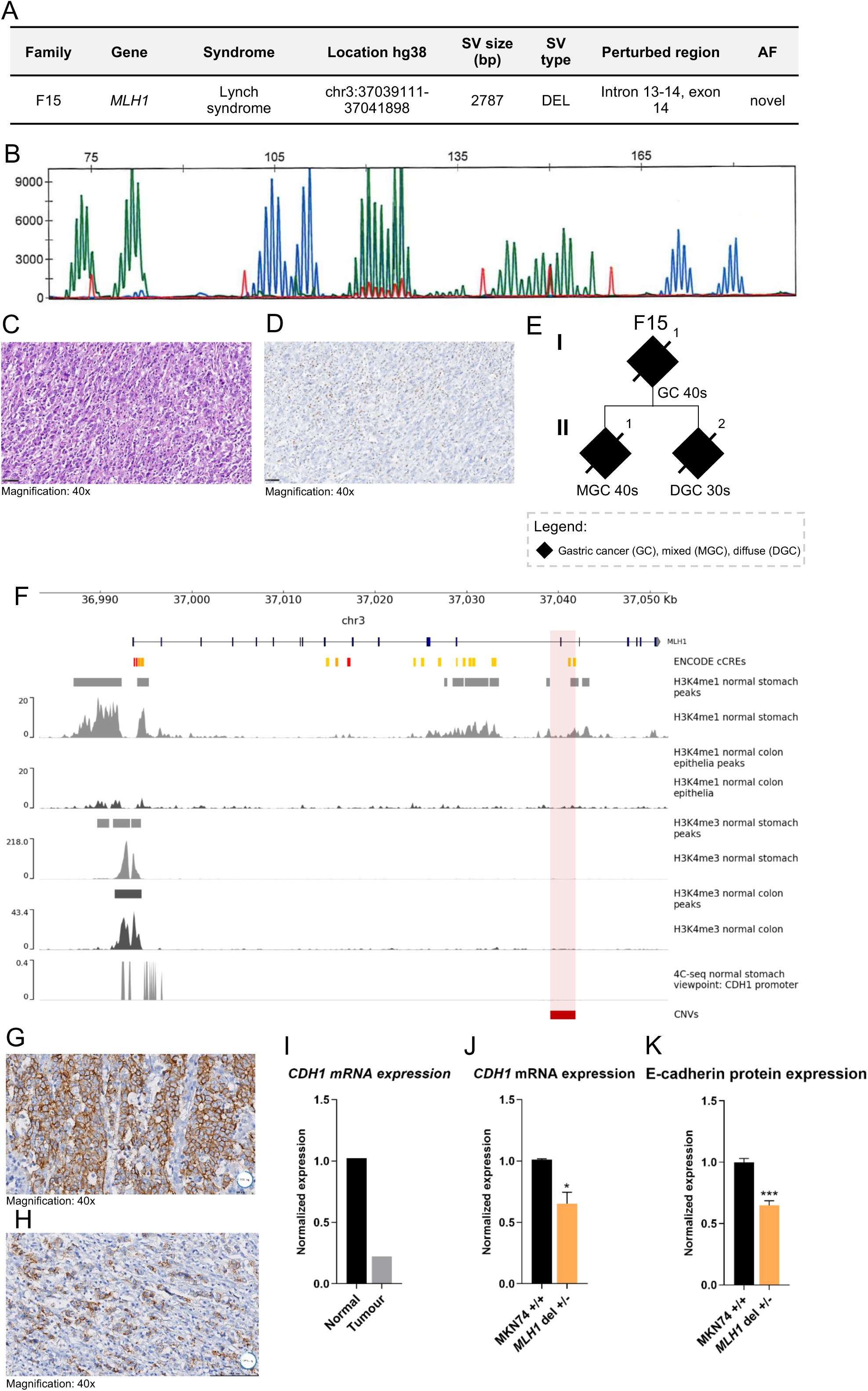
Genomic CNVs in cancer associated syndromes. A) Description of CNV identified, B) Microsatellite instability (MSI) analysis, C) Hematoxylin and eosin of tumour tissue from proband with MLH1 CNV, D) MLH1 expression of tumour tissue from proband with MLH1CNV immunohistochemistry), E) Representative family pedigree of F15. Black fill represents gastric cancer development. DGC: diffuse gastric cancer, GC: gastric cancer. F) Chromatin marks and accessibility in normal stomach and colon tissue from ENCODE, G) E-cadherin expression of tumour tissue intestinal component) from proband with MLH1 CNV immunohistochemistry), H) E-cadherin expression of tumour tissue (diffuse component) from proband with MLH1 CNV immunohistochemistry), I) CDH1 mRNA expression in the tumour, J-K) CDH1 mRNA and protein expression of edited MKN74 clones.

This family presents a proband with gastric carcinoma with lymphoid stroma and an evident diffuse component at age 40s, and another two early-onset GCs (ages 30s and 40s) in a sibling and their mother, suggesting variant segregation with disease through the maternal side (**figure 3E**). GC preponderance and lack of colorectal cancers in this family was unexpected, and raised the hypothesis that stomach-specific REs may reside within the deleted region. Indeed, the *MLH1* deletion overlaps H3K4me1 peaks, chromatin marks of active enhancers (RE3), that are present in normal stomach, but not in normal colon epithelia (**figure 3F**). RE3 lacks H3K4me3 promoter chromatin marks in both stomach and colon (**figure 3F**), suggesting this element may act as an enhancer, rather than a promoter. We resorted to 4C-seq data from normal stomach epithelia and found evidence for a long-distance interaction between *CDH1* and *MLH1* promoters (**figure 3F**), which could indicate cross-regulation between these two genes. Supporting that the *MLH1* stomach-specific RE could modulate *CDH1* expression, was the finding that E-cadherin was lost or mislocalized in the diffuse component of the proband’s tumour (**figure 3G, H**) and that *CDH1* mRNA was reduced by 80% (**figure 3I**). A CRISPR/Cas9 clone mimicking the *MLH1* heterozygous CNV triggered a reduction of 50% in the *MLH1* mRNA and protein expression levels (**supplementary figure 3 C,D**), and of 40% in the *CDH1* mRNA and protein expression levels (**figure 3J, K**).

Altogether, our results pinpoint an additional molecular mechanism, involving the germline perturbation of stomach-specific REs at the *MLH1* locus, driving somatic *CDH1*/E-cadherin downregulation and tumour MSI. This HDGC-like family may still be a Lynch syndrome family with a particular tropism to the stomach, due to a deletion of a stomach-specific RE with secondary impact in *CDH1* expression. Yet, the potential development of Lynch syndrome-associated tumours may not be disregarded in the clinic.

### Multiple germline SNVs and CNV-deletions affecting stomach-specific regulatory elements and immune-associated pathways occur in 50% of HDGC-like families

We next explored the potential of a polygenic nature for the remaining HDGC families, by evaluating the co-occurrence of variants across the genome. We searched the WGS data for ultra-rare coding deletion-CNVs and truncating SNV (AF<0.00001), prioritized by at least two callers, common in the cohort (≥6 patients, ≥24%) and absent in 13 controls from 1000genomes project. Overall, in 21 patients, we found 771 truncating SNVs and 11.860 CNVs, of which 1.547 were coding deletions. Each patient carried between 12 and 72 SNVs and 22 and 209 CNVs. When combining all genes affected by the prioritized variants in pathway analyses (**Figure 4A, supplementary figure 4**), we found a pathway-cluster that was impacted in 10 HDGC-like patients mainly presenting DGC before age 50 (8/21) (**figure 4A, B, supplementary figure 4**). This cluster included immune-associated pathways, encompassing antigen-presenting through major histocompatibility complex (MHC) class I, natural killer, tumour cell killing and T cell mediated immunity (**figure 4A, B, supplementary figure 4**). These pathways were not called for most families when considering SNVs alone, but remained represented when considering CNV only (**supplementary figure 5**), which are the main genetic event behind impairment of REs. We then asked whether deletions overlapping stomach REs support GC predisposition. By focusing on deletions overlapping accessible chromatin regions in normal stomach tissue by ATAC-seq, from 6.642 deletions, 415 overlapped accessible chromatin regions. Indeed, the same cluster of immune-associated pathways seen in HDGC-like patients based on overall CNVs and SNVs, was mimicked, despite the reduction in number of deletions overlapping stomach REs (**figure 4C, supplementary figure 6 A**). There were 39 genes recurrently impaired by deletions or truncating SNVs, and five genes recurrently deleted in their regulatory sequence, in at least three patients (**figure 6A**). We plotted these together with the monogenic events described above in Figure 6, as well as the number of patients that presented immune-pathways affected. Families F15 and F9, which presented germline *MLH1* and *CDH1*-*TANGO6* intergenic CNVs, had no other genes impaired. Six probands and F19_3 family member (invasive ductal breast cancer at age 50s) lacked variants in these pathways, mimicking the controls. Besides the *CDH3* germline CNV and SVs, F4 also displayed impaired immune-associated genes/pathways. The remaining nine probands and additional member of F19 (F19_1 with intestinal GC at 30s, F19_2 with DGC at 60s) presented between 3 and 27 deleted genes (**figure 6A**). Additionally, from 10 probands with impaired germline landscape in immune-related genes, eight had age of onset below 50 years of age (mean age of onset: 47,2 years±16,8) (**figure 6B**). While 7 individuals with deletions overlapping stomach REs had age of onset below 50 (mean age of onset: 38,4 years±8,5) from 12 individuals (**figure 6B**).

**Figure 4.**
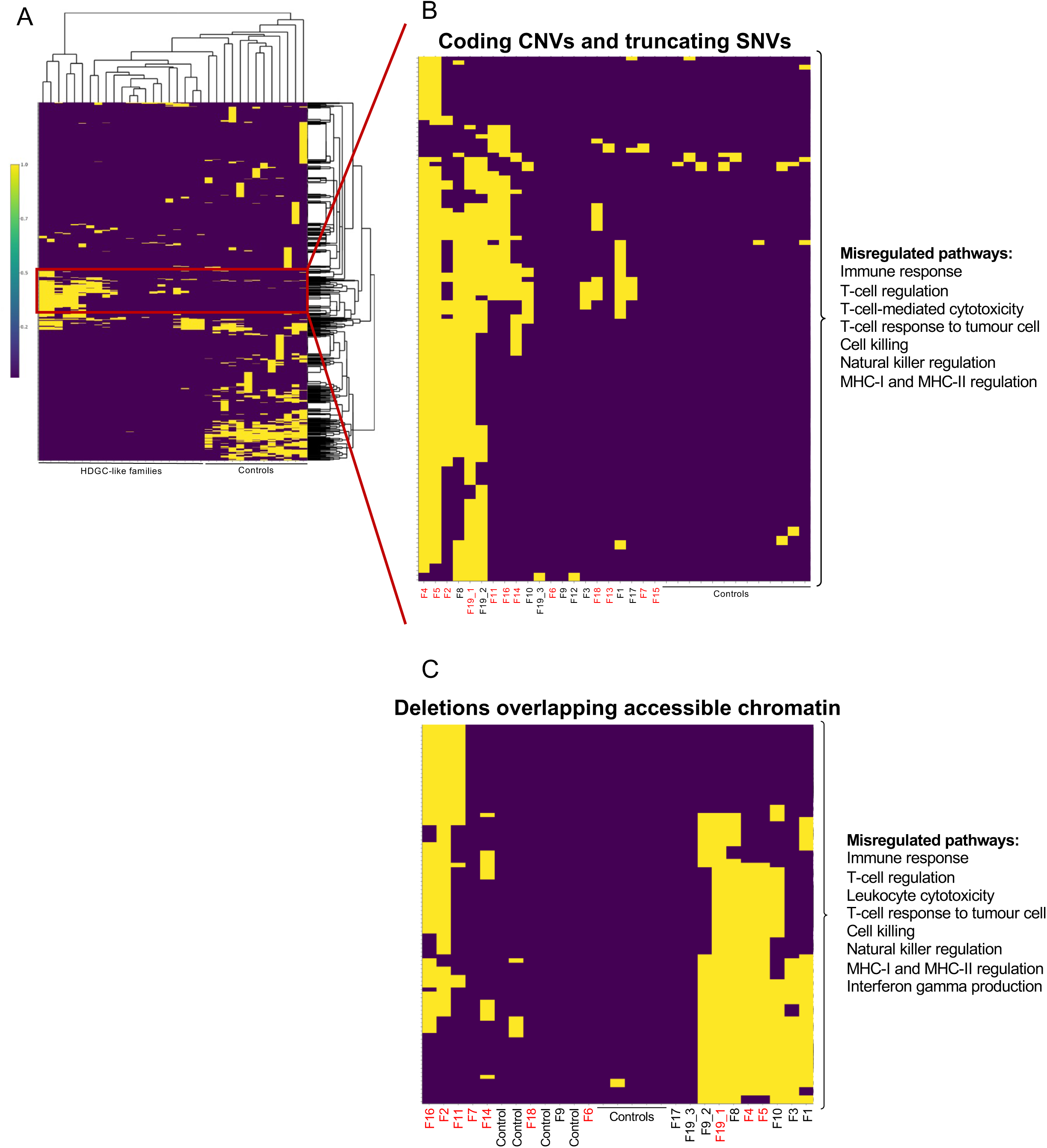
CNVs and SNVs misregulated pathways in HDGC undiagnosed patients. A) Heatmap representing all impaired pathways in HDGC undiagnosed patients, B) Cluster of immune-associated impaired pathways in HDGC undiagnosed patients and absent in control individuals, C) Cluster of CNVs overlapping accessible chromatin regions in immune-associated pathways. Red represents probands with cancer age of onset ≤50 years of age.

**Figure 5.**
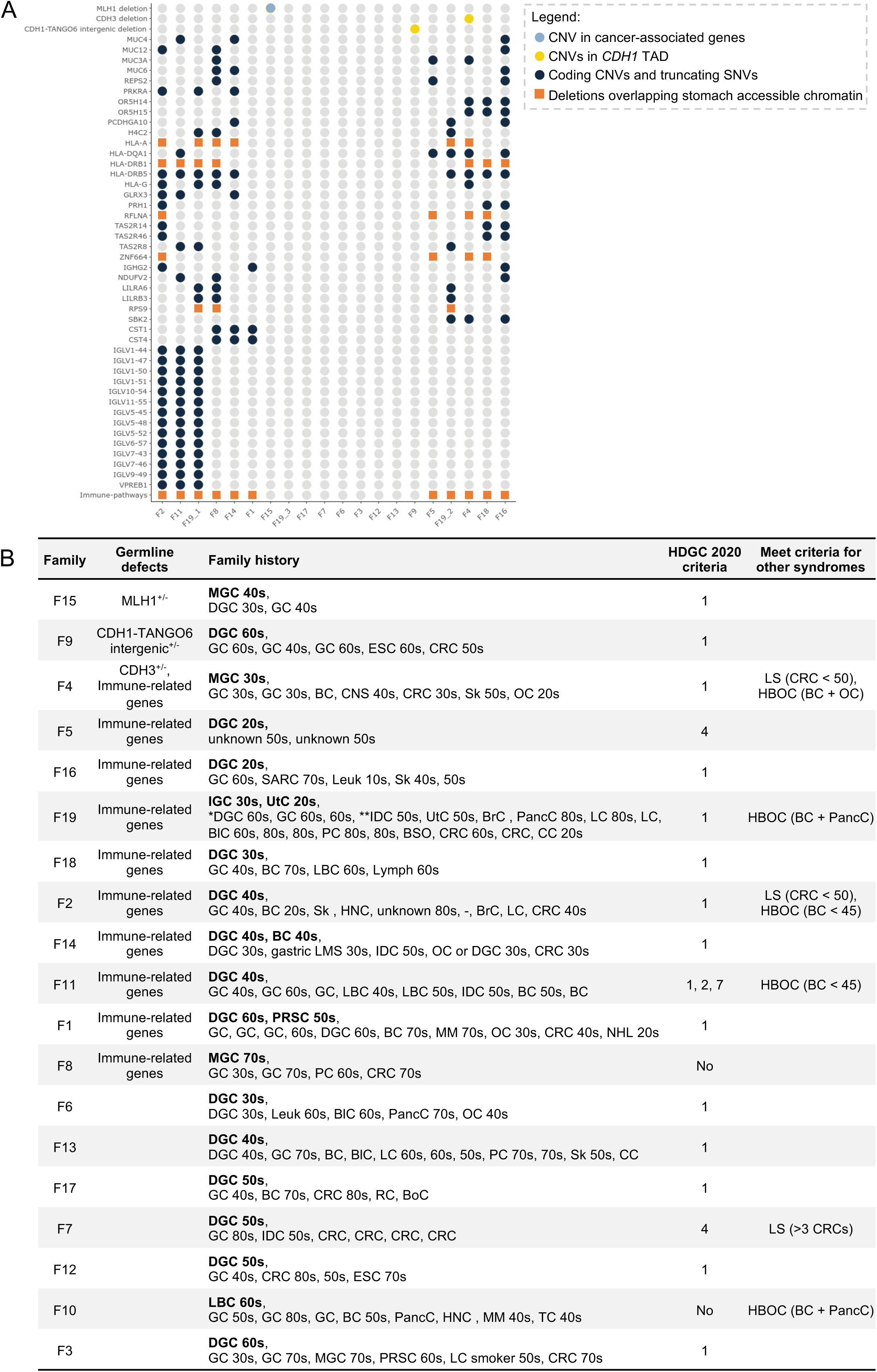
Main findings solving the missing heritability in HDGC patients. A) Heatmap representing misregulated genes in HDGC undiagnosed families, Blue represents ultra-rare (AF<0.00001) coding CNVs and truncating SNVs found from WGS sequencing of HDGC patients. Yellow represent CNVs found overlapping CDH1 TAD. Orange square represents pathways overlapping accessible chromatin in normal stomach, B) Description of the genetic defects, family history, clinical criteria and ancestry of HDGC undiagnosed families. GC: gastric cancer, DGC: diffuse gastric cancer, BC: breast cancer, LBC: lobular breast cancer, PRSS: prostate cancer, MGC: mixed-type gastric cancer, IGC: intestinal gastric cancer, IDC: invasive ductal breast cancer, CRC: colorectal cancer, EsoC: esophageal cancer, RC: renal cancer, Panc: pancreatic cancer, BldC: blader cancer, Sarc: sarcoma, OvC: ovarian cancer, Cerv: cervical cancer, UtC: uterine cancer, SkC: skin cancer, MM: melanoma, BrC: CNS: central nervous system cancer, HN: head and neck, Leuk: leukemia, NHL: non-hodkins lymphoma, Lymp: lymphoma, LC: lung cancer. In F19, * represent phenotype in individual F19_2 and ** represent phenotype in individual F19_3.

## Discussion

More than half of HDGC suspected families remain molecularly undiagnosed (HDGC-like), many of them displaying germline monoallelic *CDH1* downregulation and E-cadherin loss in the tumour (Pinheiro et al. 2010). These data pinpoint *CDH1* as the main gene triggering HDGC. Herein, we worked under the hypothesis that perturbation of REs controlling *CDH1* expression drive HDGC predisposition in HDGC-like families.

This study presents compelling evidence supporting the conclusion that cancer predisposition in 16% (3/19) of HDGC-like families within our cohort is explained by alterations occurring in the *CDH1* regulatory network, within and beyond the *CDH1* TAD. Importantly, the high frequency of HDGC-like families (10/19 – 53%) bearing ultra-rare deletions overlapping stomach REs and impairing immune-associated pathways, calls for a multigenic regulatory landscape contributing to HDGC predisposition.

The *CDH1* TAD is expected to be the most enriched region of the genome in REs modulating the *CDH1* locus (Zuin et al. 2022). Within the *CDH1* TAD, enclosing eight genes and a total of 630kb, we found a *CDH3* 20-kb deletion encompassing two noncoding hypomorphic and dose-dependent enhancers controlling *CDH1* expression, which likely represent a novel biological mechanism of HDGC predisposition in family F4. The *CDH1*/E-cadherin loss in the diffuse component of the tumour further argues towards the role of this deletion in HDGC predisposition. The combined deletion of two *CDH1* enhancers, each contributing to half *CDH1* expression loss, represents a classical mechanism of enhancer-driven expression control in *CDH1-*expressing tissues (Stemmler et al. 2005; Will et al. 2017). In this family, this heterozygous *CDH3*-CNV produces an expression loss similar to that induced by a heterozygous coding truncating *CDH1* variants and is expected to lead to germline *CDH1* monoallelic expression, which would explain the occurrence of GC with a diffuse component in the proband, her brother and mother. Indeed, CD44-Cre/*Cdh1^fl^*^/fl^ mice develop stage 1 mixed-type gastric cancer (Decourtye-Espiard and Guilford 2023), correlating well with a tumour with mixed-type histology reduced *CDH1* expression. Although we could not assess the impact of the *CDH3* inversions, likely inherited from the paternal side, these are more likely to impact *CDH3* expression rather than *CDH1* expression, as the enhancers are orientation-independent. In fact, we found that inversion of the 20kb deleted region in F4, does not impact *CDH1* expression (*data not shown*). The compound heterozygous state found for *CDH3* (allele 1: deletion + allele 2: inversions) in F4 proband, leading to nearly complete loss of *CDH3* expression, supports the tumour spectrum found in this family, which goes beyond the classical HDGC disease spectrum (Garcia-Pelaez et al. 2023). In contrast to normal stomach epithelia, which does not express *CDH3*, ovary and skin do (Cao et al. 2020), which may explain why biallelic loss of *CDH3* may predispose to the ovarian and skin tumours observed in the family. Even if only one of the *CDH3* alleles has been inherited by other affected family members, it may still cause cancer predisposition, as combined loss of function occurs for both *CDH1* and *CDH3* (allele 1: deletion) with a single allele being targeted. Furthermore, the central nervous system tumour occurrence correlates well with the forebrain enhancer activity observed for the RE1 and RE2 contained within the *CDH3*-CNV. *CDH3* loss of expression through combined deletion of the *CDH3* promoter, exon 1 and 2, and/or an inversion of the 5’-region of *CDH3* may also trigger ectodermal dysplasia, ectrodactyly and macular dystrophy syndrome or congenital hypotrichosis with juvenile macular dystrophy syndrome, both autosomal recessive (OMIM ID 601553). However, as far as we know, these diseases have not been reported for this family.

The *CDH1*-*TANGO6* intergenic sequence also encompasses a noncoding hypomorphic enhancer, but homozygous deletion triggers loss of only half *CDH1* expression. In addition, the mean cancer age of onset in family F9 (63.7 years of age) is higher than what is usually found for HDGC families (42±12,16 years of age) (Garcia-Pelaez et al. 2023). Taken together, a third inactivation event may be required for GC development in F9 carriers, which would take more time to occur. This 3-hit hypothesis would in part explain the later cancer onset in this family. Deletion of this intergenic region in combination with a 3’-coding deletion of the *CDH1* has been shown to trigger extremely early GC onset and high disease penetrance (São José et al. 2023a). That deletion, in contrast to the current one, strongly reduced *CDH1* expression and led to re-wiring of the 3D chromatin structure, impairing the transcriptome, immune, proliferation and adhesion-associated pathways, as compared to a deletion of *CDH1* alone (São José et al. 2023a). Thus, we propose that deletion of REs lying in the *CDH1*-*TANGO6* intergenic sequence occurring in both families, modulates *CDH1* expression and predisposes carriers to GC development. This likely occurs later in life in F9, because the *CDH1* coding region remains wild-type (São José et al. 2023a).

In a knowledge-driven approach, we next explored rare CNVs and SNVs in cancer-syndrome-associated genes and found a germline *MLH1* deletion, that has not been previously identified in Lynch syndrome families to date, according to the InSiGHT database. Although *MLH1* loss of function predisposes individuals to Lynch syndrome, preferentially increasing the risk of colorectal and endometrial cancers, intestinal-type GC may also occur, with an estimated risk of 13% at age 80 (Møller et al. 2018). In family F15, the clinical history is restricted to GC, but the proband’s tumour presents not only diffuse, but also intestinal histology, which is consistent with *MLH1* expression loss and MSI (Møller et al. 2018). Further supporting the hypothesis that GC predisposition is associated with *MLH1* in this family, somatic *MLH1* promoter methylation, a hallmark of MSI in sporadic colorectal and GCs, was negative in the proband’s tumour (Shen et al. 2018). Our data raised the hypothesis that *CDH1* expression disruption was involved not only in generating the diffuse component of this tumour, but also in shifting the *MLH1*-related tumour spectrum to the stomach. Indeed, not only were *CDH1*/E-cadherin mRNA and protein found to be downregulated in the tumour, but also the deleted sequence encompassed enhancer chromatin marks present in normal stomach tissue and absent from normal colon mucosa. Also, an interaction found by 4C-seq between *CDH1* and *MLH1* promoters further supported a cross-regulation between both genes. Rare inter-chromosomal interactions have been observed among neighbouring chromosomal territories, such as CISTR-ACT in chromosome 12 and SOX9 in chromosome 17 (Quinodoz et al. 2018; Milad Mokhtaridoost et al. 2024). Altogether, these data indicate that the *MLH1* exon 13 sequence, and its intronic vicinity, act as a tissue-specific RE, controlling the expression of *CDH1*, specifically in the stomach. This regulatory module and the loss of *MLH1* expression, as a consequence of deleting an out-of-frame exon, would concomitantly trigger MSI in the emerging GCs. To our knowledge, this is the first report associating a *MLH1* exon 13 deletion with *CDH1* loss, and its associated stomach-specific REs with GC predisposition.

The novel genetic alterations described above, as potential causes for three HDGC-like families, do not affect the structure of the *CDH1* locus, but rather involve *CDH1* enhancers located either within the *CDH1* TAD or located in another chromosome. In all instances, the downregulation of *CDH1* occurs due to transcriptional impairment caused by deletion of enhancers for which long distance interactions have been identified herein. Similar mechanisms have been described for several genes predisposing to developmental disorders (Will et al. 2017).

It is also worth mentioning that probands from HDGC-like families F4 (*CDH3* CNVs and SVs) and F15 (*MLH1* CNV) present mixed GC with a diffuse component, rather than pure DGC, which suggests that impairment of *CDH1* REs outside of the *CDH1* locus may have wider impacts than the impairment of the *CDH1* locus alone. This becomes evident not only in the different histology of GCs, but also in the diversity of cancers in family F4.

Given the lack of obvious variants in the remaining 16 families, we explored the germline regulatory landscape of HDGC undiagnosed families. The germline regulatory landscape of undiagnosed families encompasses mainly downregulation of immune-associated pathways. A specific cluster was found for younger affected individuals, with downregulation of pathways associated with MHC I and natural killer immunity, absent in healthy controls, which are known to play a role in GC immune evasion (Shen et al. 2005; Na et al. 2021). Interestingly, earlier onset DGC patients display a higher number of deletions in genome-wide open chromatin regions in normal stomach tissue, compared to patients with older cancer onset. These “younger-onset”-related regions encompass immune-affected pathways, recapitulating the germline regulatory landscape and pinpointing stomach REs as possible genetic drivers. Altogether, the observed immune impairment in HDGC families may result in lower immunological surveillance in early stages of intramucosal signet-ring cell carcinoma *foci*, characteristic of HDGC tumours. Indeed, variants in immune surveillance genes, such as MHC class I, have been suggested to play a role in intramucosal signet-ring cell carcinoma *foci* (Decourtye-Espiard and Guilford 2023), yet their consolidation is requires additional research. Interestingly, families F15 and F9 lack additional alterations to the *MLH1* CNV and the *CDH1*-*TANGO6* intergenic CNVs, suggesting these were sufficient to trigger the HDGC phenotypes. Indeed, F15 (*MLH1* CNV) tumour presents high degree of inflammation, highlighting that the lack of germline defects in immune-related pathways allows the immune system to recognize tumor cells. In this particular case, the F15 proband would benefit from immunotherapy treatment (Chao et al. 2021). In contrast, family F4 displays impaired immune-associated genes/pathways as well as the *CDH3* CNV and SVs.

Immune-associated genes and several mucins were also found impaired in other HDGC-like families. Supporting the role of *MUC4* in this context is the reduced expression described in poorly differentiated GC or signet-ring cell carcinoma (Tamura et al. 2012), also characteristic of HDGC (Carneiro et al. 1995; van der Post et al. 2016).

Downregulation of the stomach-specific *MUC6* gene (Babu et al. 2006) was found to be associated with advanced stages of GC and poor prognosis (Shi and Xi 2021), supporting an association with HDGC. Indeed, homozygous deletion of *MUC6* causes spontaneous GC development with accumulation of CD45^+^ T-cells in normal and tumour tissue (Arai et al. 2024). Impairment of T-cell regulatory mechanisms and MHC-related immune surveillance in the germline with co-occurrence of mucins reduced expression may pinpoint a polygenic mechanism for GC development in HDGC families.

Altogether, impairment of immune- and mucins-associated pathways/genes may constitute a germline genetic background capable of regulating HDGC development in the presence of a primary unknown triggering mechanism.

Taken together, our data resolves the missing heritability in 3 out of 19 HDGC undiagnosed families, corresponding to 16% of the entire cohort, and pinpointed novel mechanisms of HDGC predisposition related with *CDH1* REs as regulators of *CDH1*/E-cadherin expression. The finding of a causal mechanism promotes re-engagement with the family, genetic counselling and disease risk management of individuals at risk. The germline landscape of younger DGC patients includes downregulation of immune-associated pathways and several mucins, which may play a role in immune surveillance and DGC development when triggered by a still unknown mechanism.

## Data Availability

All data produced in the present study are available upon reasonable request to the authors.

## Acknowledgments

This work received funding from: (1) the European Union’s Horizon 2020 research and innovation programme under grant agreement No 779257 (Solve-RD: H2020-SC1-2017-Single-Stage-RTD), which provided Ph.D. salaries to JG-P; (2) The European Regional Development Fund (ERDF) through the COMPETE 2020-Operacional Programme for Competitiveness and Internationalisation (POCI), Portugal 2020, and by Portuguese funds through Fundacao para a Ciencia e Tecnologia (FCT)/Ministerio da Ciencia, Tecnologia e Inovação (3DChroMe: PTDC/BTM-TEC/30164/2017, LEGOH: PTDC/BTM-TEC/6706/2020, DGCoding: 2023.11994.PEX); (3) FCT with salary support for PhD studentships to CSJ (Ref. SFRH/BD/140796/2018), MF (Ref. 2020.05763.BD) and SL (Ref. 2020.05773.BD). Further, this study has received funding from the European Union’s Horizon Europe Research and Innovation action under the Grant Agreement n° 101095483 (PREVENTABLE). Views and opinions expressed are however those of the author(s) only and do not necessarily reflect those of the European Union or The Health and Digital Executive Agency (HaDEA). Neither the European Union nor the granting authority can be held responsible for them.

This work was supported by: (1) Doctoral Programme in Biomedicine - Faculty of Medicine, University of Porto, to CSJ and JG-P; (2) Doctoral Programme in Computational Sciences, Faculty of Sciences, University of Porto, to MF; (3) Doctoral Programme on Cellular and Molecular Biotechnology Applied to Health Sciences, School of Medicine and Biomedical Sciences, University of Porto, Portugal, to SL.

## Author Contributions

Carla Oliveira: supervision, Project administration, funding acquisition; Celina S. José and Carla Oliveira: conceptualization, formal analysis; José García-Pelaez, Janine Sanz, Lilian Cordova, Pardeep Kourah: acquisition of clinical and pathological data; Irene Gullo: pathological analysis; Celina S. José, Ana Pedro, Ana André, Silvana Lobo: *in vitro* experimental work; Marta Ferreira and Celina S. José: bioinformatic analysis; Celina S. José: integration of genetics, clinical and functional data; Celina S. José, Fiona Puntieri and Juliane Glaser: *in vivo* experimental work; All authors: writing, review, editing and approval of the manuscript.

